# Confirmed forecasts for the expansion of the COVID-19 epidemic in the largest Brazilian City

**DOI:** 10.1101/2021.02.08.21251290

**Authors:** Sergio Celaschi

## Abstract

**Objective:** A SEIR compartmental model was previously selected to estimate future outcomes to the dynamics of the Covid-19 epidemic breakout in Brazil.

**Method:** Compartments for individuals vaccinated, and prevalent SARS-Cov-2 variants were not included. A time-dependent incidence weight on the reproductive basic number accounted for Non Pharmaceutical Interventions (NPI). A first series of published data from March 1^st^ to May 8, 2020 was used to adjust all model parameters aiming to forecast one year of evolutionary outbreak. The cohort study was set as a city population-based analysis.

**Analysis:** The population-based sample, 25,366 during the study period, was the number of confirmed cases on exposed individuals. The analysis was applied to predict the consequences of holding for posterior NPI releases, and indicates the appearance of a second wave starting last quarter of 2020.

**Findings:** By March ^1st^2021, the number of confirmed cases was predicted to reach 0.47Million (0.24-0.78), and fatalities would account for 21 thousand (12-33), 5 to 95% CRI. A second series of data published from May 9, 2020 to March 1^st^, 2021 confirms the forecasts previously reported for the evolution of infected people and fatalities.

**Novelty:** By March 1^st^ 2021, the number of confirmed cases reached 527,710 (12% bellow predicted average of accumulated cases), and fatalities accounted for 18,769 (10% above the accumulated average of estimated fatalities). After March 1^st^, new peaks on reported numbers of daily new infected and new fatalities appeared as a combined result to the appearance of the prevalent SARS-CoV-2 P1 variant, and the increased number of vaccinated individuals.

## 1. Introduction

COVID-19 was first reported in Brazil in February 2020. One semester later, the country became one of the worst affected globally. After six months from the first reported case, the number of confirmed cases and deaths crossed 3.9 million and 120 thousand, respectively. Facts that question the availability of public health care for a major fraction of society [1, 2]. As of October 2020, all parts of the world were, to varying degrees, impacted by the COVID-19 epidemic, with more than 40 million cases and 1.1 million deaths reported globally [3]. This work aims to confirm previous forecasts reported on May, 2020 [4] for the expansion of the COVID-19 epidemic in S. Paulo, the most populated Brazilian city with its 12.2 million inhabitants. The city of S. Paulo was selected once the first patient in Brazil was tested positive there. Since then, by April 25, 2021, were officially confirmed in Brazil 14.4 million cases with 391 thousand deaths, and in S. Paulo city 704,989 cases, and 26,642 deaths. Before the beginning of vaccination, the worldwide response to the pandemic has been the introduction of Non Pharmaceutical Interventions (NPIs) as mitigation policies.

Regarding predictions, epidemiological models are commonly stochastic, diffusive-spatial, network based, with heterogeneous sub-populations (meta-population approaches) [5, 6, 7, 8, 9]. However, the parameters of compartmental models are more directly related to and interpretable as physical processes [10, 11, 12]. On the other hand, deterministic models impose restrictive analysis, once the dynamics of the host population and the virus are not deterministic. The population has free will, and the virus undergoes “random” mutations [13, 14]. The primarily intent of this work was to build a simple epidemiological and compartmental model to predict and confirm the main results for the basic dynamics of the Covid-19 epidemic breakout in the city of S. Paulo, Brazil. Compartments for individuals vaccinated, and prevalent SARS-Cov-2 variants were not included to the model [15]. The real-time transmissibility of an infectious disease is often characterized by the instantaneous reproduction *R(t)* defined as the expected number of secondary infections caused by an infector within a short time window. Equivalently, *R(t)* can be expressed as the transmission rate *β(t)* divided by the rate *γ*_*o*_ into which infected people recover or die. Mitigation policies aim to control the outbreak reducing the *R(t)* value. In this work, a time-varying incidence weight on the basic reproductive number R_o_ was adopted to account for external influences as an elementary empirical approach. A first series of official published data from March 1st to May 8, 2020 [4] was used to adjust the model parameters aiming to forecast one year of the COVID-19 evolutionary outbreak. The analysis was applied to predict the consequences of NPI enforcements follow by progressive releases, and indicates the appearance of a second epidemic wave. A second series of official published data from May 9, 2020 to March 1^st^, 2021 confirms the forecasts previously reported for the evolution of infected people and fatalities associated to this epidemic outbreak in the city of S. Paulo.

## 2. Compartmental model and Methodology

The compartmental SEIR model (Fig. 1) represents one of the most adopted mathematical models to characterize an epidemic dynamics, and to predict possible contagion scenarios. The SEIR model can be useful to assess the effectiveness of various measures, such as social distancing, lock-down, mask wearing, closing public areas, among others. The model is based on a series of dynamic ordinary differential equations that consider the amount of the population subject to contagion, the trend over time of individuals who get exposed, recover after infection, and the individuals who unfortunately die. The coefficients of the equations represent the ratios of variation over time of different compartments, i.e., susceptible, exposed, infected, recovered, and dead. These coefficients have often been considered as constants. However, as constants they cannot take into account external influences, such as the NPIs, as social distancing, mask wearing, lockdowns, closed public places, or the possible change in health conditions of infected individuals due to pharmacological development. In this approach, a time-dependent model parameter was introduced. It was assumed the infection rate time-dependent, considering that the number of contacts between people, during NPIs, changes proportionally to their overall mobility. Enforced NPIs in the city of S. Paulo reduced the growth of infected after the third week (March 22, 2020). Additionally, the fatality rate μ was set constant, which is an oversimplification not supported by data. In Figure 1, the susceptible (S) is part of the population that could be potentially subjected to the infection. The exposed (E) is the fraction of the population that has been infected but is not infectious: it can be called a latent phase. The infective (I) represents the infective hosts after the latent period. The recovered (R) accounts for the fraction of hosts after healing, R hosts are not reintroduced into the susceptible category assuming they became immune to the disease during the study period. The fatality (F) is the dead part of the population N considered in this cohort study, *S(t) + E(t) + I(t) + R(t) + F(t) = N*. As can be noticed in this figure, compartments for individuals vaccinated, and prevalent SARS-CoV-2 variants were not included to the model.

**Figure 1.**
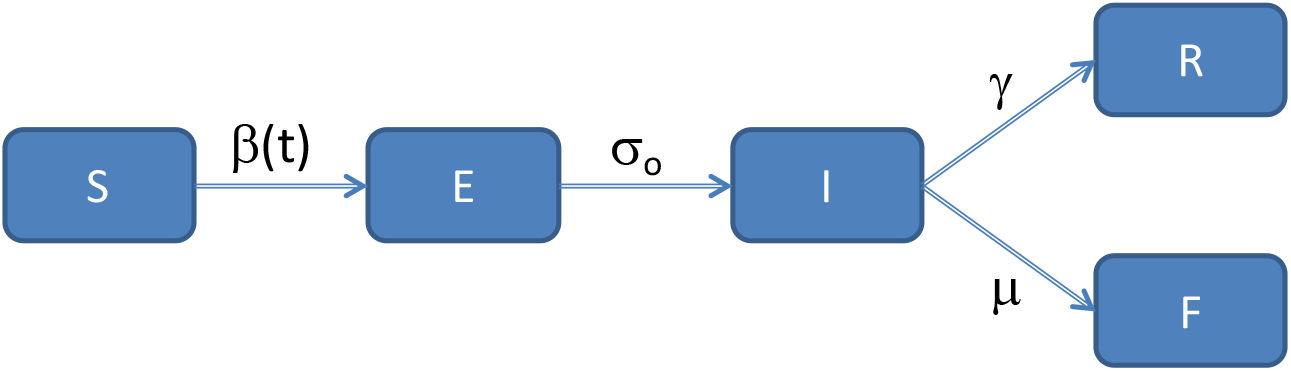
Compartmental epidemiologic model used in this cohort study. The coefficients represent the ratios of variation of different compartments, i.e., susceptible, exposed, infected, recovered, and dead.

The research methodology employed in this work is presented as a flowchart in Fig. 2. The largest populated city in Brazil (S. Paulo) was selected in this cohort study. The basic concepts applied to the selected SEIR model are available elsewhere [4].

**Figure 2.**
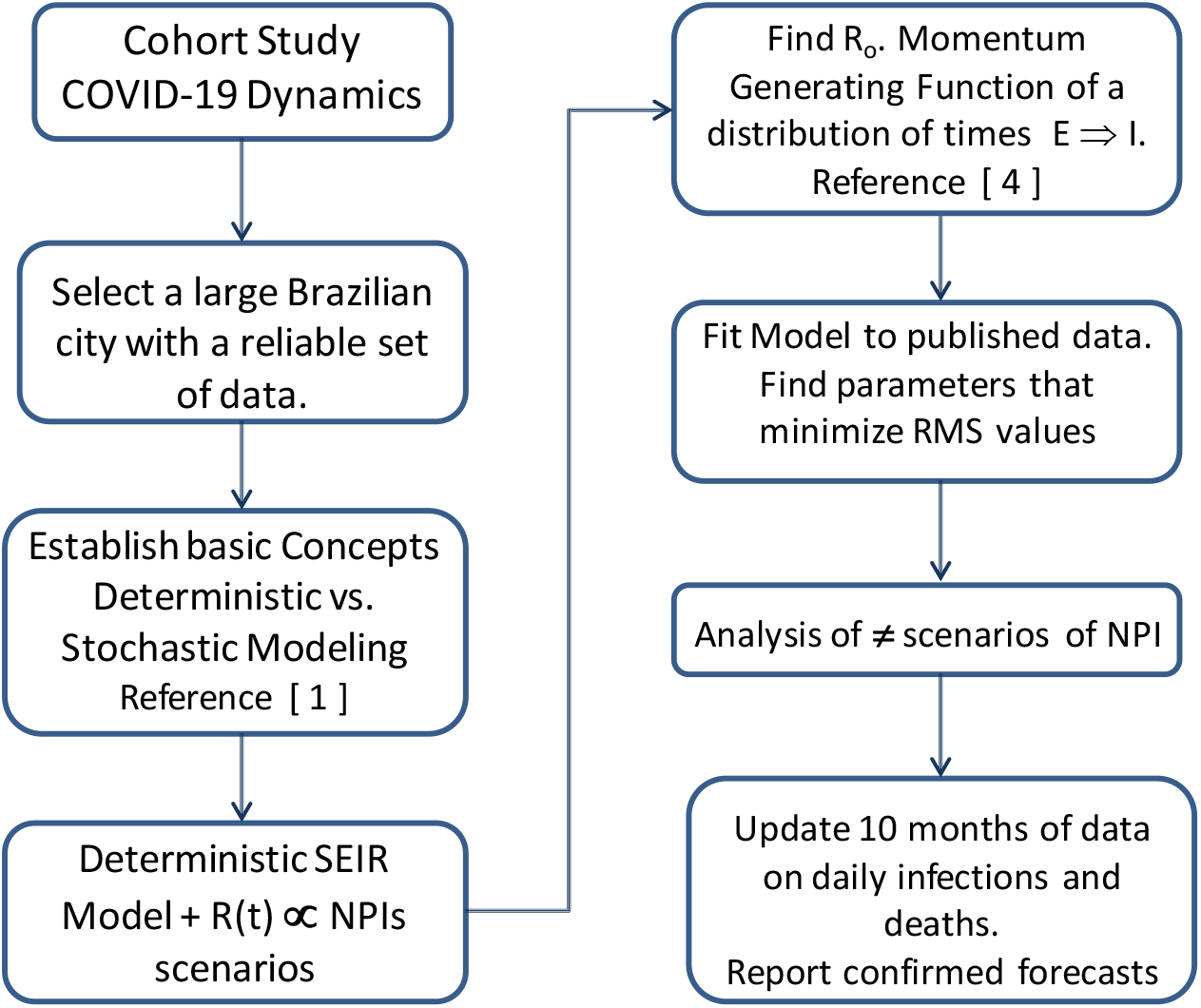
Flowchart for the research methodology applied to this work.

A first series of published data from March 1st to May 8, 2020 was initially considered to estimate the epidemiological parameters that govern the selected compartmental model dynamics, and to adjust model parameters aiming to forecast one year of the COVID-19 evolutionary outbreak. The model provides predictions of the time series of infected individuals and fatalities in the studied area. Simulations of midi-term scenarios of the epidemic outbreak were done dependent on the level of NPIs enforcement and release. Forecasts showed how such a policy alters the pattern of contamination, and suggested the existence of a second epidemic wave.

A key parameter in deterministic transmission models is the basic reproductive number *R*_*o*_, which is quantified by both, the pathogen and the particular population in which it circulates. Thus, a single pathogen, like the SARS-CoV-2, will have different *R*_*o*_ values depending on the characteristics and transmission dynamics of the population experiencing the outbreak. The methodology to estimate *R*_*o*_ *= 2.53 ± 0.05* was presented in a previous publication [4]. Accordingly, *R(t)=ψ(t).R*_*o*_ becomes dependent on the NPI policy *ψ(t)*. The analysis of a time-varying reproductive number *R(t)* is applied to forecast the consequences of enforce, maintain and release the NPIs over specific periods of time. By May 2020, during the first part of this study, without enough data on asymptomatic hosts, the percentage of symptomatic host’s ξ_o_ was arbitrarily set at 50%. Later on, applying statistical analysis on published and reliable data, the percentage value of non-symptomatic and symptomatic hosts were estimated to be respectively (54±9) % and (46±9) % [14].

Figure 3 illustrates a simulation of daily infected hosts as the result of the enforcement and the progressive realize of NPIs during the period of study. The NPI ending time releases was selected to best fit the reported data on infected hosts. Surprisingly or not, the model reported on May, 2020 suggests the existence of a second outbreak wave of Covid-19 characterized by the presence of a new peak of daily infected appearing months after the first one. The number of infected individuals was estimated to be lower compared to the first outbreak. This is a consequence of the partial reduction in the initial number *N* of susceptible individuals.

**Figure 3.**
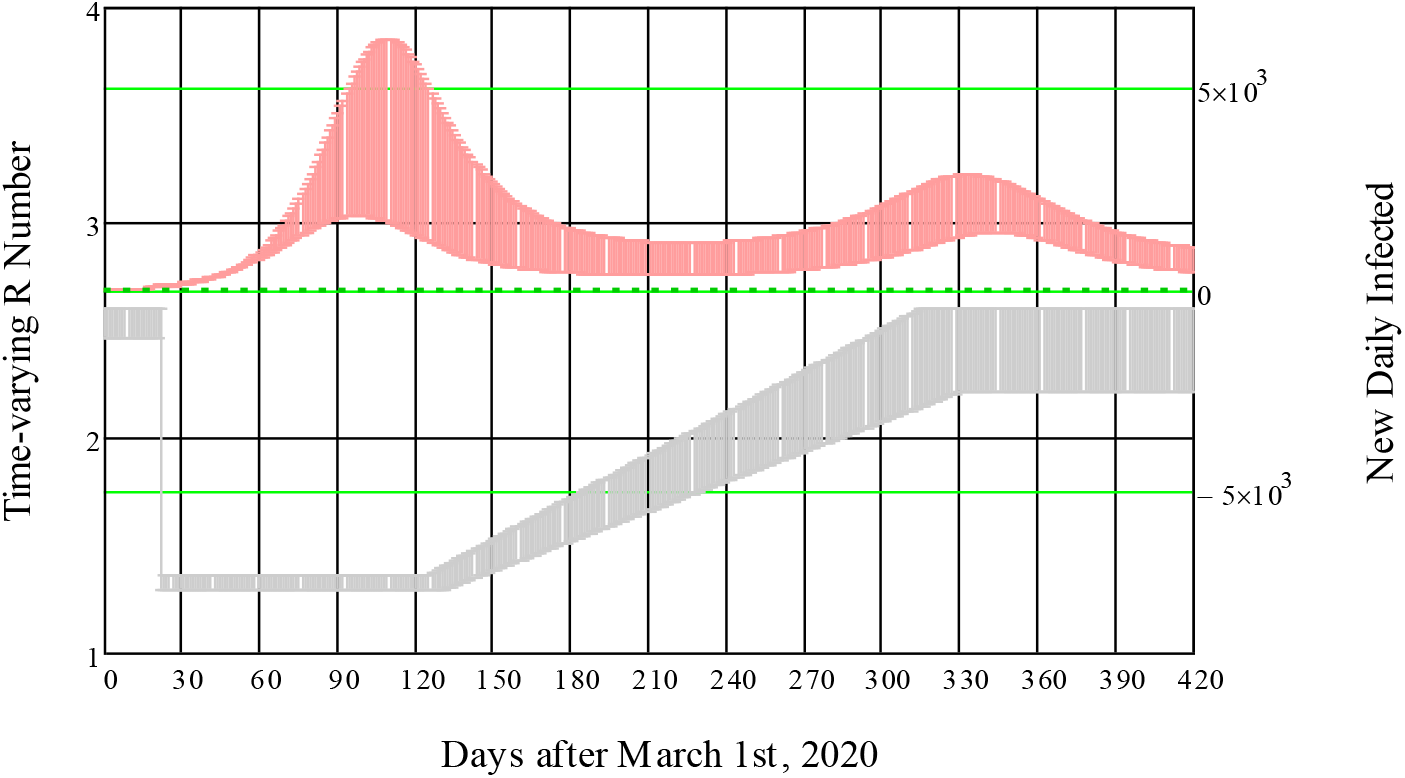
Long term prediction (red shaded area) of daily infected hosts (right vertical axis) as a consequence of proposed variations of R(t) (left vertical axis - gray shaded area) modeling the NPI after March 22^nd^, 2020 in the city of S. Paulo. The values of the SEIR parameters are: Ro = 2.53 ± 0.05, N_average_=8.5×10^5^ (5×10^5^ <N<14×10^5^), β_o_ = 0.980±0.012, γ_o_ = 0.387±0.018, ψ _o_ = 0.525, μ = 0.015±0.001, σ_o_ = 0.5, ξ_o_ = 0.5.

## 3. Discussion and Forecasts

The fittings to data on accumulated infected individuals, and fatalities by the selected SEIR model are shown in Fig. 4. A first series of official published data from March 1st to May 8, 2020 was previously used to adjust the model parameters aiming to forecast about one year of the COVID-19 evolutionary outbreak (Fig. 4a). The reported fitting values are: *5×10*^*5*^ *< N <14×10*^*5*^, *R*_*o*_ *= 2.53±0.05, β*_*o*_ *= 0.980±0.012, γo = 0.387±0.018, ψ* _*o*_*= 0.525, μ = 0.015±0.001, σ*_*o*_ *= 0.5*. The cohort study was set as a city population-based analysis. The population-based sample, 25,366 during the initial study period, was the number of confirmed cases on exposed individuals. In the same period, the accumulated fatalities accounted for 2,110 confirmed deaths.

**Figure 4.**
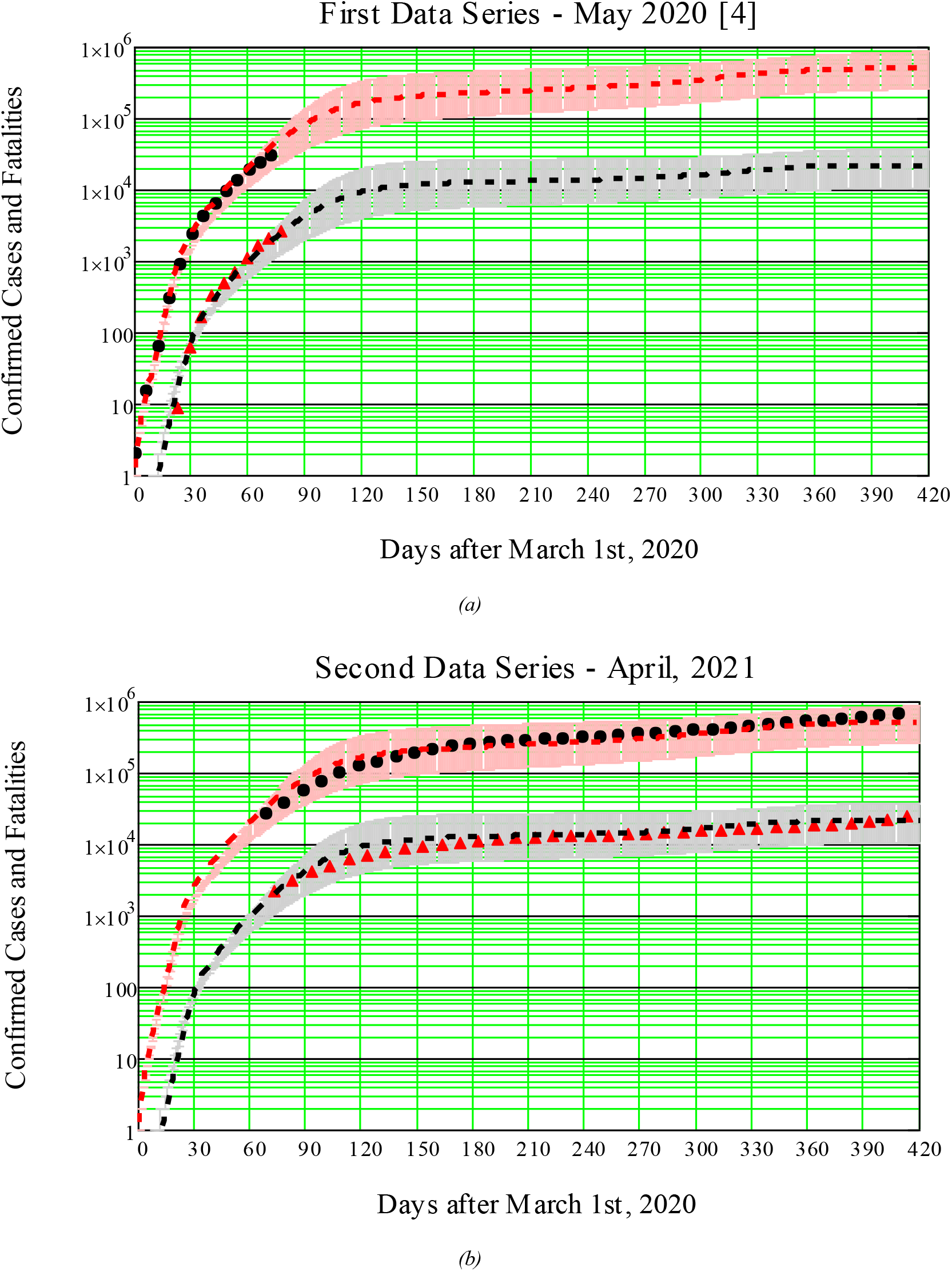
(a) Primary data on confirmed cases and fatalities are shown by solid black circles, and red triangles respectively. Shadow areas account for the 5-95% CRI on model’s parameters. The fitting mean values to model are: N = 8.5×10^5^, R_o_ = 2.53, β_o_ = 0.980, γ = 0.387, μ = 0.015, ψ _o_= 0.525, σ_o_ = 0.50, and t = 22 days. (b) Second series of official published data from May 9, 2020 to April 24, 2021 on confirmed cases and fatalities are shown by solid black circles, and red triangles respectively.

A second series of official published data [16] from May 9, 2020 to April 24, 2021 shown in Fig. 4b, confirms the forecasts reported previously on May 2020 for the evolution of infected people and fatalities associated to this epidemic outbreak in the city of Sao Paulo, Brazil. The average fitting value to confirmed cases (logarithmic scale) for the whole study period presents standard deviation SD = 0.07 and root mean square RMS = 0.08. By March 1^st^ 2021, the official number of confirmed cases reached 509,193 an increase of 20 folds on the population-based sample (25,366), with 12% deviation above the predicted average value. The reported fatalities accounted for 18,769 (10% below the average of predicted deaths). In short, the previous forecasts reported on May, 2020 for the expansion of the COVID-19 epidemic in the most populated Brazilian city with its 12.2 million inhabitants are confirmed by the epidemiological SEIR model as shown in Fig. 3b. During the period March 2^nd^ to April 25, 2021, the last day reported, new sharp peaks appears on the reported numbers of daily new infected and new fatalities. These are caused by a combined result to the appearance of the prevalent SARS-CoV-2 P1 variant, and the increased number of vaccinated individual, as commented later.

The data ratio, fatalities to confirmed cases in S. Paulo, present a temporal evolution that is not linear neither monotonic (Fig. 5), as previously reported for the whole Brazilian country [14]. The non-zero ratio of deaths/(confirmed cases) by COVID-19 (open red circles), begins two weeks after the first reported case, rises monotonically, after roughly seven weeks reaches a maximum of 0.085, and drops continuously to a ratio around 0.04. Possible explanations are many: Increased number of tests and reports available after the first weeks of the epidemic outbreak raises the number of confirmed cases; ICU improved medical procedures; among others. COVID-19 was detected in Brazil in the 9th EPI week of 2020, and testing procedures for the SARS-CoV-2 virus was effectively included in the surveillance four weeks later. Such a time dependent ratio was not included in the compartmental model. The fatality rate μ was set constant throughout the study period, which leads to a monotonic behavior as shown in Fig.4a by the black dash-and-dot line. This simplification leads, for the same time period, to an overestimation of the infected host as presented in Fig. 4, and degrades the values of SD and RMS. Adding to model a time-varying fatality rate μ(t) by data fitting to the first 90 days after March 1st, 2020 (Fig. 5b) would further improve the values of SD and RMS to 0.05 and 0.06 respectively.

**Figure 5.**
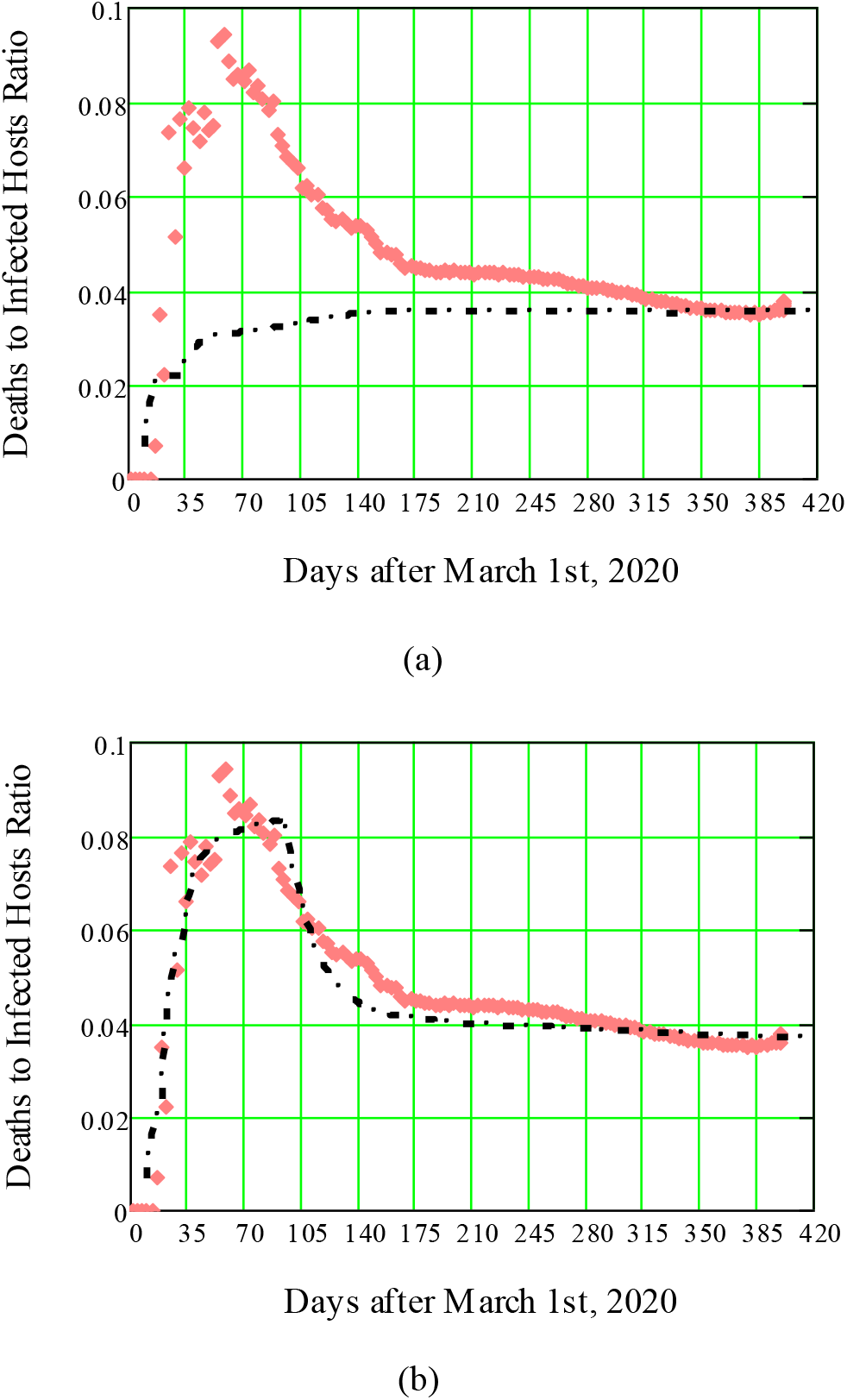
Ratio of deaths to confirmed cases (open red diamonds) as a function of time. (a) An assumed constant fatality rate in the compartmental model leads to a monotonic behavior in disagreement to data (shown by the black dash-and-dot line). (b) A time-varying fatality rate μ(t) (not included in the previous report) obtained by data fitting to the first 90 days after March 1st, 2020, would further improve the SD and RMS values.

Novel SARS-CoV-2 lineage P.1 first identified in Manaus has been associated with potentially higher transmission rates, and now it is widespread across all Brazilian regions [17, 18]. On the other hand, a preliminary study conducted by Institute Butantan [19], suggests the vaccination in Brazil by CoronaVac can neutralize variants P.1 and P.2 of SARS-CoV-2. P.1 was first worldwide reported on January 10, 2021 after been detected in four travelers from Brazil at Haneda airport in Tokyo. It is circulating widely in Brazil and is considered a “variant of concern” because it is more transmissible. Epidemiologists believe P.1 is one of the major causes for recent jump in COVID-19 cases and deaths, alongside the relaxing of mobility restrictions in and after the holiday season and the slow pace of vaccination. Since February 15, in a period of 12 weeks, the variant P.1 of Covid-19, which emerged in Manaus, increased its representativeness in the cases of the Sã o Paulo city from 0% to 91%. The estimate was based on virus samples taken from patients at Hospital Sã o Paulo, and confirms the prevalent spread of the new lineage [20]. As the result of this explosive spread, during the period March 2^nd^ to April 25, 2021, the last day reported in this study, sharp peaks appears on the reported numbers of daily new infected and new fatalities, as presented in Figure 5. These sharp peaks are caused by the combined result to the appearance of the prevalent SARS-CoV-2 P1 variant, and the increased number of vaccinated individual. As mentioned previously, compartments for individuals vaccinated, and prevalent SARS-Cov-2 variants were not included to this simple model, and their combined effects were not predicted.

Regarding data on daily new cases and fatalities, due to a large data scattering, a 7 days moving average was applied to smooth data. Part of scatter data is due to the unreported cases and deaths during weekends and holydays, which accumulates large values on coming working days. Assuming the variation of R(t) as already modeled for the NPI enforcement and posterior progressive release (Fig.3), a comparison to the reported numbers of daily new cases and fatalities is presents in Fig. 5 in comparison to model predictions. Reducing data scattering bring the data points closer to average values. The low agreement between data and model during the first outbreak wave is, in part, caused by the assumption of a constant fatality rate.

**Figure 5.**
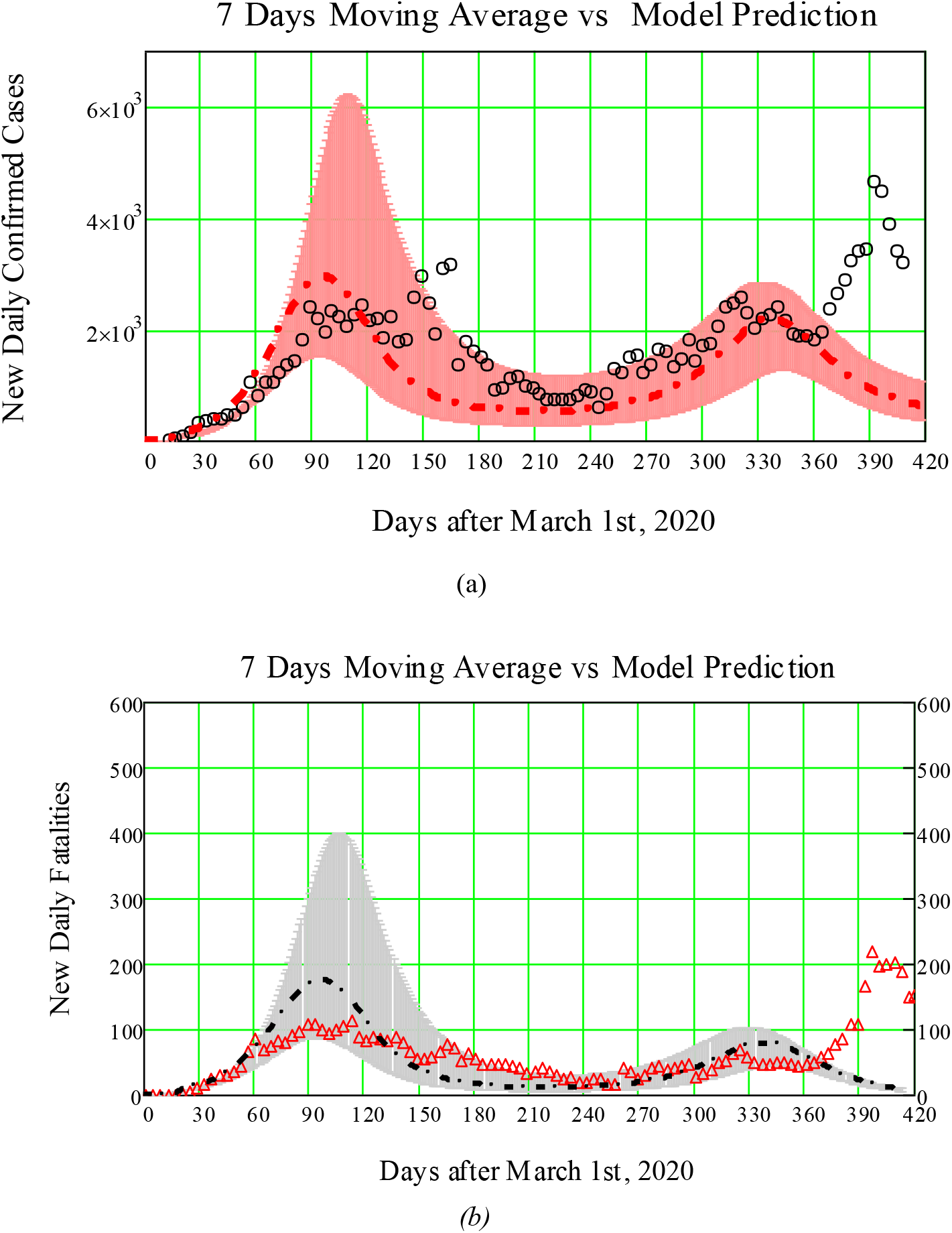
Reported (a) daily new cases (solid red circles) and (b) fatalities (solid black triangles) as a function of time compared to model predictions. The presence of a second wave becomes evident by published data and model predictions shown as shaded areas.

This basic assumption leads to an overestimation of the first estimated peak of confirmed cases and fatalities. The presence of a second wave becomes evident by the data and model predictions presented by Fig. 5a and 5b. Surprisingly or not, the previous publication forecasted the presence of a second peak months before its arrival. As mentioned before, the predicted number of infected individuals within the second peak was estimated to be lower compared to the first one. The sharp peaks on confirmed cases and deaths at the end of the study period are the combined result to the appearance of the prevalent SARS-CoV-2 P1 variant, and the increased number of vaccinated individuals.

## 4. Conclusions

This work confirms the forecasts previously reported on May, 2020 for the evolution of infected people, fatalities, and the appearance of a second wave starting last quarter of 2020 associated to the epidemic outbreak in S. Paulo, the largest Brazilian city [4]. The cohort study was set as a city population-based analysis in a sample of 25,366 confirmed COVID-19 cases on exposed symptomatic individuals. The analysis was applied to predict the consequences of the progressive NPI releases previously enforced during the first semester of 2020. An updated series of published data from May 9, 2020 to March 1^st^, 2021 confirms the forecasts previously reported for the evolution of infected people and fatalities associated to this epidemic outbreak. By March 1^st^ 2021, the official number of confirmed cases reached 527,710 an increase of 20 folds over the population-based sample (25,366), with 12 % deviation bellow the predicted average value. The reported fatalities accounted for 18,769 (10% above the average of predicted deaths). During the period March 2^nd^ to April 25, 2021, as a result of the explosive spread of the prevalent SARS-CoV-2 P1 variant, and the increased number of vaccinated individuals, new sharp peaks appear on the reported numbers of daily new infected and new fatalities. The simple SEIR model employed in this study did not include compartments for vaccinated individuals, and prevalent SARS-Cov-2 variants. As a result, their combined effects were not predicted.

## Data Availability

All available data in the manuscript was previously published by the Brazilian government.

http://www.saopaulo.sp.gov.br/wp-content/uploads/2021/01/20210131_dados_covid_municipios_sp.csv

## 5. Acknowledgements

The author acknowledges the financial grant by FUNTTEL - Financial grant # 01.16.0053.01 FINEP/MCTI, Brazilian Ministry of Science, Communications, and Innovation.

## 8. Declaration of Data availability

All data presented in this study is available in article [16].

## 9. Ethical Approval

The manuscript does not contain experiments on animals and humans; hence ethical permission not required.

